# Graph Convolutional Neural Networks for Histological Classification of Pancreatic Cancer

**DOI:** 10.1101/2022.01.26.22269832

**Authors:** Weiyi Wu, Xiaoying Liu, Robert B. Hamilton, Arief A. Suriawinata, Saeed Hassanpour

## Abstract

Pancreatic ductal adenocarcinoma has some of the worst prognostic outcomes among various cancer types. Detection of histologic patterns of pancreatic tumors is essential to predict prognosis and decide about the treatment for patients. This histologic classification can have a large degree of variability even among expert pathologists. This study proposes a graph convolutional network-based deep learning model to detect aggressive adenocarcinoma and less aggressive pancreatic tumors from benign cases. Our model uses a convolutional neural network to extract detailed information from every small region in a whole-slide image. Then, we use a graph architecture to aggregate the extracted features from these regions and their positional information to capture the whole-slide level structure and make the final prediction. We evaluated our model on an independent test set and achieved an F1 score of 0.85 for detecting neoplastic cells and ductal adenocarcinoma, significantly outperforming other baseline methods. If validated in prospective studies, this approach has a great potential to assist pathologists in identifying adenocarcinoma and other types of pancreatic tumors in clinical settings.

## INTRODUCTION

Pancreatic ductal adenocarcinoma (PDAC) is an aggressive type of cancer derived from the epithelial cells that make up the ducts of the pancreas. PDAC ranks firmly last among all cancer types in terms of worst prognostic outcomes ^1^, and its incidence and mortality rates have continued to increase for decades in Unites States ^2–5^. According to one study ^6^, from 1990 to 2017, the number of cases and deaths worldwide identified as related to pancreatic carcinoma doubled. It is estimated that there will be 60,430 pancreatic cancer cases and 48,220 deaths caused by pancreatic cancer in the United States in 2021 ^7^. Furthermore, PDAC is expected to become the second leading cause of cancer death by 2030 ^8^. Therefore, preventive measures, screening, and surveillance are becoming increasingly important for pancreatic cancer.

PDAC makes up the overwhelming majority of pancreatic malignant tumors and is derived from the ductules of the exocrine pancreas, which carry digestive enzymes and other secretions from the exocrine pancreas to the lumen of the small bowel. PDAC has a variable histologic appearance, ranging from high-grade lesions with necrosis and marked cellular atypia to bland, “foamy” infiltrative glands in a highly fibrotic stroma. Inflammation may be prominent, subtle, or essentially absent. The islet cells of the pancreas also have a neoplastic counterpart, which is the pancreatic neuroendocrine tumor, or PanNET. PanNET, while prognostically much more favorable, is also histologically diverse and may mimic benign inflammatory condition (e.g., islet aggregation in chronic pancreatitis) or PDAC.

Endoscopic ultrasound-guided fine-needle aspiration (EUS-FNA) and EUS guided fine-needle biopsy (EUS-FNB) ^9^ have become the primary diagnostic methodology employed in the evaluation of pancreatic mass lesions. These methods are the least invasive means of procuring tissue for diagnosis currently available, as they can be performed via endoscopy. However, they have significant limitations in terms of tissue fragmentation, crush artifact, and the overall quantity of tissue procured for diagnosis.

Considering these limitations, the severity of pancreatic carcinoma, and the enormous morbidity of pancreatectomy, it is imperative that the diagnostic utility of EUS-FNA/FNB be maximized. PDAC is a histologically diverse malignant neoplasm with numerous known patterns, including several which can mimic neuroendocrine tumors (that typically have a much better prognosis) and various non-neoplastic pancreatic lesions. Although there exists a set of guidelines for the classification of pancreatic tumors from the Papanicolau Society of Cytopathology (see **Table 1**), some cases can be ambiguous even to the trained eyes of pathologists. The low tissue volume of FNA/FNB procedures may increase the frequency of cases that do not receive a definitive diagnosis (i.e., cases labeled as “Atypical” or “Suspicious”). Therefore, not only radiologists and oncologists but also pathologists may incorrectly identify some benign tumors or pseudo-tumor cells for malignant tumors ^10–12^.

**Table 1.**
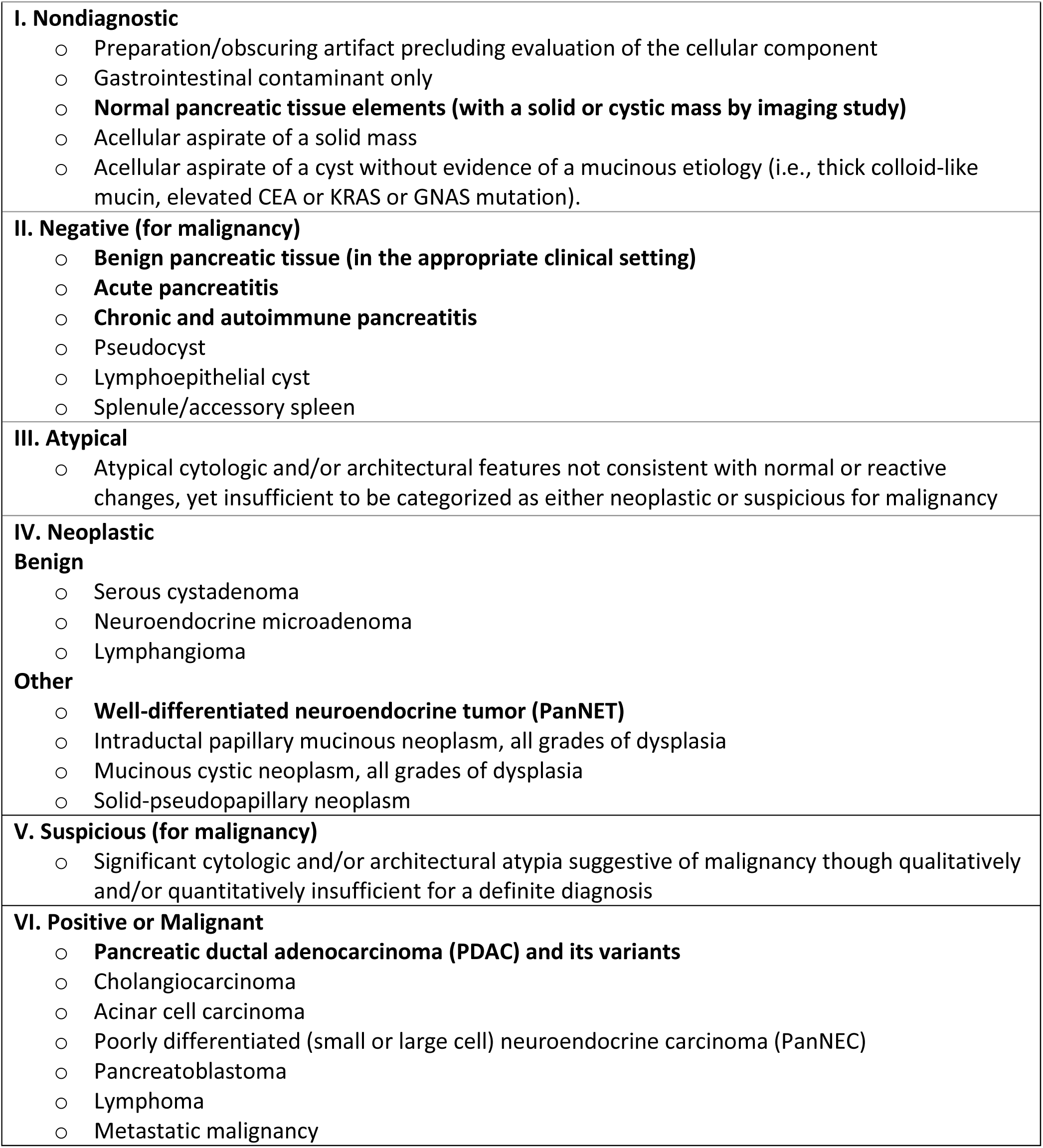
Classification criteria modified from Papanicolaou Society of Cytopathology System for Reporting Pancreaticobiliary Cytology ^13^. The bold cases are those classes that are included in this study.

|  |
| --- |
| **I. Nondiagnostic**<br>○ Preparation/obscuring artifact precluding evaluation of the cellular component<br>○ Gastrointestinal contaminant only<br>○ **Normal pancreatic tissue elements (with a solid or cystic mass by imaging study)**<br>○ Acellular aspirate of a solid mass<br>○ Acellular aspirate of a cyst without evidence of a mucinous etiology (i.e., thick colloid-like mucin, elevated CEA or KRAS or GNAS mutation). |
| **II. Negative (for malignancy)**<br>○ **Benign pancreatic tissue (in the appropriate clinical setting)**<br>○ **Acute pancreatitis**<br>○ **Chronic and autoimmune pancreatitis**<br>○ Pseudocyst<br>○ Lymphoepithelial cyst<br>○ Splenule/accessory spleen |
| **III. Atypical**<br>○ Atypical cytologic and/or architectural features not consistent with normal or reactive changes, yet insufficient to be categorized as either neoplastic or suspicious for malignancy |
| **IV. Neoplastic**<br>**Benign**<br>○ Serous cystadenoma<br>○ Neuroendocrine microadenoma<br>○ Lymphangioma<br>**Other**<br>○ **Well-differentiated neuroendocrine tumor (PanNET)**<br>○ Intraductal papillary mucinous neoplasm, all grades of dysplasia<br>○ Mucinous cystic neoplasm, all grades of dysplasia<br>○ Solid-pseudopapillary neoplasm |
| **V. Suspicious (for malignancy)**<br>○ Significant cytologic and/or architectural atypia suggestive of malignancy though qualitatively and/or quantitatively insufficient for a definite diagnosis |
| **VI. Positive or Malignant**<br>○ **Pancreatic ductal adenocarcinoma (PDAC) and its variants**<br>○ Cholangiocarcinoma<br>○ Acinar cell carcinoma<br>○ Poorly differentiated (small or large cell) neuroendocrine carcinoma (PanNEC)<br>○ Pancreatoblastoma<br>○ Lymphoma<br>○ Metastatic malignancy |

Our study proposes an automatic and accurate method based on graph convolutional neural networks to detect pancreatic neuroendocrine tumors (PanNET) and PDAC on digitized histology slides. Furthermore, such an approach can assist pathologists with reviewing the slides by generating additional diagnostic information for consideration, such as the location of cells suspicious for malignancy or neoplasia in a pancreatic tissue specimen.

## DATA COLLECTION AND ANNOTATION

In this study, we focused on classifying the two most common pancreatic tumors, PDAC and PanNET, in combination with a benign “control” group. To develop and evaluate our model to classify these pancreatic tumor patterns, we collected 143 digitized formalin-fixed paraffin-embedded (FFPE) hematoxylin and eosin (H&E)-stained whole-slide images from Dartmouth-Hitchcock Medical Center (DHMC). The EUS-FNA/EUS-FNB cell block slides were digitized using an Aperio AT2 scanner (Leica Biosystems, Nussloch, Germany) at 20× resolution (0.5 μm/pixel) at DHMC. These slides were identified using structured cytopathology diagnosis data from the laboratory information management system (LIMS). In addition, a full-text pathology report search was performed for further disambiguation of cases within each class. To assure the quality of the slides and their labels in our dataset, they were independently reviewed by the expert pathologists involved in our study (R.H. & X.L.) for concordance with the identified diagnosis. Our pathologists assessed all slides identified as negative, positive, and those with neoplastic cells present based on LIMS structured data and pathology reports. A manual review of report text was employed for cases of unusual histologic appearance or any other irregularity concerning the identified diagnosis versus the slide’s appearance.

Our classification criteria for this dataset are described in **Table 1**. Of note, cases without viable tumors (e.g., entirely necrotic) were excluded from the dataset. The “Positive” cases in our dataset only include PDAC. At the same time, lymphomas and other malignancies were excluded due to the availability of an insufficient number of those cases for training at our institution. The “neoplastic cells” class in our dataset is represented by neuroendocrine tumors, which exclude neuroendocrine carcinomas. Also, rarer tumors such as solid pseudopapillary tumors were excluded from this class. In addition, we opted to exclude cystic lesions, such as mucinous cystic neoplasm and intraductal papillary mucinous neoplasm, in the neoplastic category because the diagnosis of these cases often relies heavily on cyst fluid chemistry studies and clinical information, which our proposed neural network does not consider. The negative class contains normal cells as well as blood, fibrin, mild atypia associated with inflammation, leukocytes, benign gastric or duodenal epithelium due to procedural artifact. “Atypical” and “Suspicious” cases were also excluded due to lack of definitive diagnosis and inter-observer variability. Rarely, some cell blocks from such cases were included in the negative category if they met one of two criteria: (1) The original pathology report described them as negative or unremarkable; (2) They were cleared in blinded review by our senior expert cytopathologist (X.L.).

The digitized slides in our dataset were partially annotated by our domain expert pathologists (R.H. and X.L.) to indicate the pancreatic cancer subtypes and their locations on the slides. As such, the neoplastic and positive regions are annotated using the polygon annotation feature in the Automated Slide Analysis Platform (ASAP) ^14^, a fast-viewer and annotation tool for high-resolution histology images. These annotations are used as reference standards for developing and evaluating our patch classification models. The distribution of the annotated images is shown in **Table 2**. Any disagreements in annotations were resolved through joint discussions among annotators and further consultation with our senior expert pathologist (X.L.). We randomly partitioned negative slides and the 52 annotated slides into a training set, a validation set, and a test set for the patch level classification.

**Table 2.** Distribution of our dataset and its annotations

|  | Annotated Slides |  | All Slides (Annotated+ Unannotated) |  |  |
| --- | --- | --- | --- | --- | --- |
|  | Neoplastic | Positive | Negative | Neoplastic | Positive |
| Training | 13 | 18 | 32 | 28 | 30 |
| Validation | 2 | 3 | 5 | 4 | 5 |
| Test | 7 | 9 | 16 | 11 | 12 |
| Total | 22 | 30 | 53 | 43 | 47 |

Of note, all whole-slides (annotated and unannotated) were reviewed and classified by our expert pathologists (R.H. & X.L.), and whole-slide labels were established according to consensus opinions between X.L. and R.H., and the original diagnosis in the clinical report. Slides on which agreement could not be reached were excluded from our dataset. All whole-slide images were randomly partitioned into the training, validation, and test sets and used for whole-slide inference.

## METHODS

Given the large size of high-resolution whole-slide histology images and the memory capacity of currently available computer hardware, it was not feasible to directly train a model on whole-slide images. Therefore, we use a sliding window strategy to extract small fixed-size (224×224 pixel) patches from the whole-slide images. To analyze and classify a whole-slide image, our pipeline has two parts, (1) a deep convolutional neural network to extract high dimensional features from patches extracted from a whole-slide image and (2) a graph convolutional neural network to aggregate high dimensional features and positional information to make the whole-slide inference. As a result, this pipeline allows us to analyze high-resolution images with feasible memory and computational resource.

### Deep Convolutional Neural Network

Deep neural networks have been proved a powerful tool in computer vision and are increasingly applied in medical image analysis ^15–17^. In the histologic image inference domain, deep convolutional neural networks are applied as a backbone for whole-slide image analysis and classification ^18,19^. This study uses a residual neural network ^20^ to extract the image features. The whole-slide images are usually high resolution, from 0.25μm/pixel to 1μm/pixel. Because of this high resolution and hardware memory limitations, it is not feasible to directly extract features from whole-slide images without downsampling. However, by the downsampling of whole-slide images, we may lose critical histological features for classification. Therefore, to train our deep residual neural network with achievable memory and computational resource requirements, we use a sliding-window strategy to generate small, fixed-size patches from the whole-slide images. The labels of these small patches depend on whether they include the annotated regions of interest by pathologists. Using this sliding-window approach, we generated 3,091 neoplastic patches, 6,275 positive patches, and 9,4633 negative patches in the training set. We then trained multiple ResNet models with different numbers of layers, including 18, 50, and 101 layers. Among these, the ResNet-18 model performed the best in the patch level classification on our validation set. Therefore, we used the ResNet-18 model trained on the augmented annotated training set as our feature extractor. In the training process of the feature extractor, the ResNet-18 model used the extracted tissue patches as inputs and outputted the predicted class probabilities for each patch. All the layers in this model are initialized with He initialization ^21^. We trained the ResNet-18 model for sixty epochs with an initial learning rate of 0.001 and decayed the learning rate by a factor of 0.9 to the power of the number of epochs.

### Graph Neural Network

Using the ResNet-18 model, we can extract features from patches and get the predictions at the patch level. To infer the whole-slide image labels, we developed a novel method based on a graph neural network to aggregate the patch level information extracted by our patch level ResNet-18 model for the whole-slide level inference. In recent years, graph neural networks, such as graph convolutional networks, have gained massive success in analyzing data with non-regular structures, such as social networks and protein networks ^22,23^. A graph convolutional network uses convolutional layers to aggregate the neighbor nodes’ information and has achieved state-of-the-art performance on graph classification benchmarks such as Citeseer, Cora, Pubmed, and NELL ^24–27^. Some recent work has proposed utilizing graph convolutional networks to make the whole-slide inference ^28^. This method leverages the pre-trained ResNet-50 model on ImageNet ^29^ to extract features from patches in a whole-slide image. Although this approach’s overall architecture is similar to ours, our approach is different in constructing the graph and extracting the patch-level features. Of note, instead of the ResNet-50 model pre-trained on ImageNet, our feature extractor uses ResNet-18 architecture and is trained on labeled pancreas patches from annotations. In our approach, the whole-slide images are viewed as graphs. Fixed-size patches and their extracted features are considered nodes and node features, respectively. We use the patches’ positional information and features extracted by our patch classification model to construct graphs from whole-slide images, which we named Slide2Graph method and described below.

### Slide2Graph for Whole-Slide Inference

#### Graph Construction

An overview of our graph construction pipeline is shown in **Figure 1**. To construct the computational graphs for whole-slide images, we first use a framework developed by our group for slide preprocessing to automatically remove background and extract tissue segments at a 10× (1μm/pixel) magnification level ^30^. Then, tissue images are divided into 224×224 pixel fixed-size patches, and the coordinates of patches are saved. We removed the last fully connected (FC) and the SoftMax layers from our patch-level ResNet-18 classifier and utilized the rest of our trained ResNet-18 to extract 512-dimensional feature vectors from each fixed-size patch. Each 224×224 pixel fixed-size patch image extracted from a whole-slide image is viewed as a node in the computational graph, and its 512-dimensional feature is used as the node feature. We used the previously saved positional information of patches or nodes to add edges in the computation graphs. For each node, we used the KD-Tree algorithm ^31^ to search its four nearest nodes in the Euclidean space and then add edges between nodes which were weighted by the nodes’ Euclidean distance. As a result, we converted the whole-slide images into computational graphs. We used graph convolutional networks (GCNs), which leverage all patches’ spatial and positional information to aggregate the local patch features and make the whole-slide image-level inference. Of note, in comparison to previous aggregation methods, this method incorporates the positional information of each patch and the global structure of the whole-slide image.

**Figure 1.**
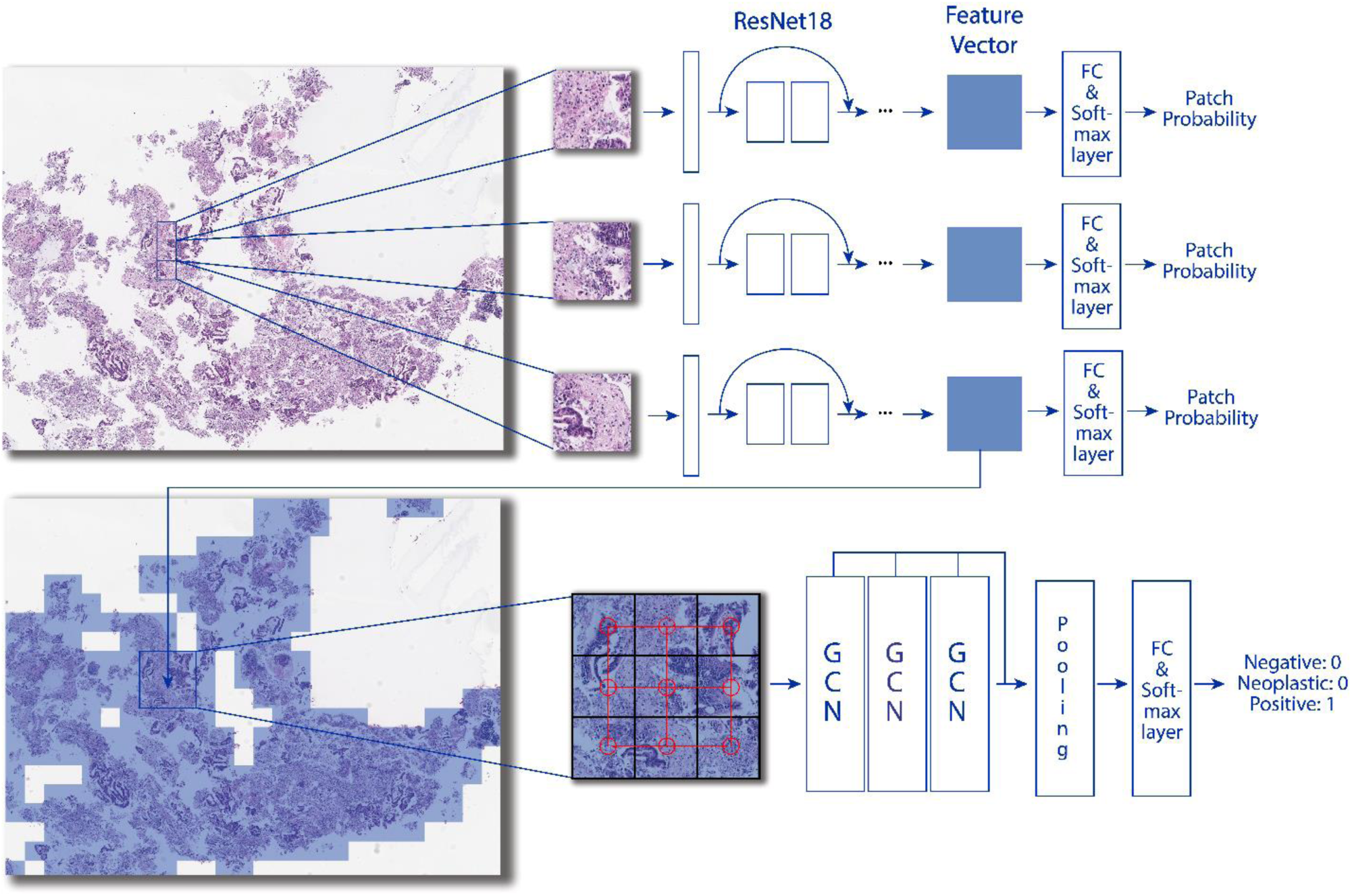
Overview of our Slide2Graph classification pipeline. After background removal, fixed-size patches are extracted from whole-slide images using the sliding-window method. A ResNet-18 model was trained on the extracted patches from annotated whole-slide images and then used to extract histology features of patches. The features and positional information of patches were used to construct a computational graph for whole-slide inferencing.

#### Graph Model Architecture

We modify the graph model proposed by Zhang et al. ^32^ and use self-attention global pooling layers ^33^ as our graph model architecture. The overall model structure is shown in **Figure 2**. After constructing the graph, we apply three graph convolutional or GCN layers to update the node features. Therefore, every node contains the information of its neighborhoods. We concatenate the outputs of every GCN layer and then utilize the self-attention pooling layer to select the top 50% highly weighted nodes that determine the class of a graph. We run a global mean pooling and maximum pooling on these top 50% nodes and concatenate them. Finally, a fully connected layer and a SoftMax layer take the features matrices and output the predicted whole-slide class probabilities. We trained this model for 200 epochs with an initial learning rate of 0.001 and cosine annealing.

**Figure 2.**
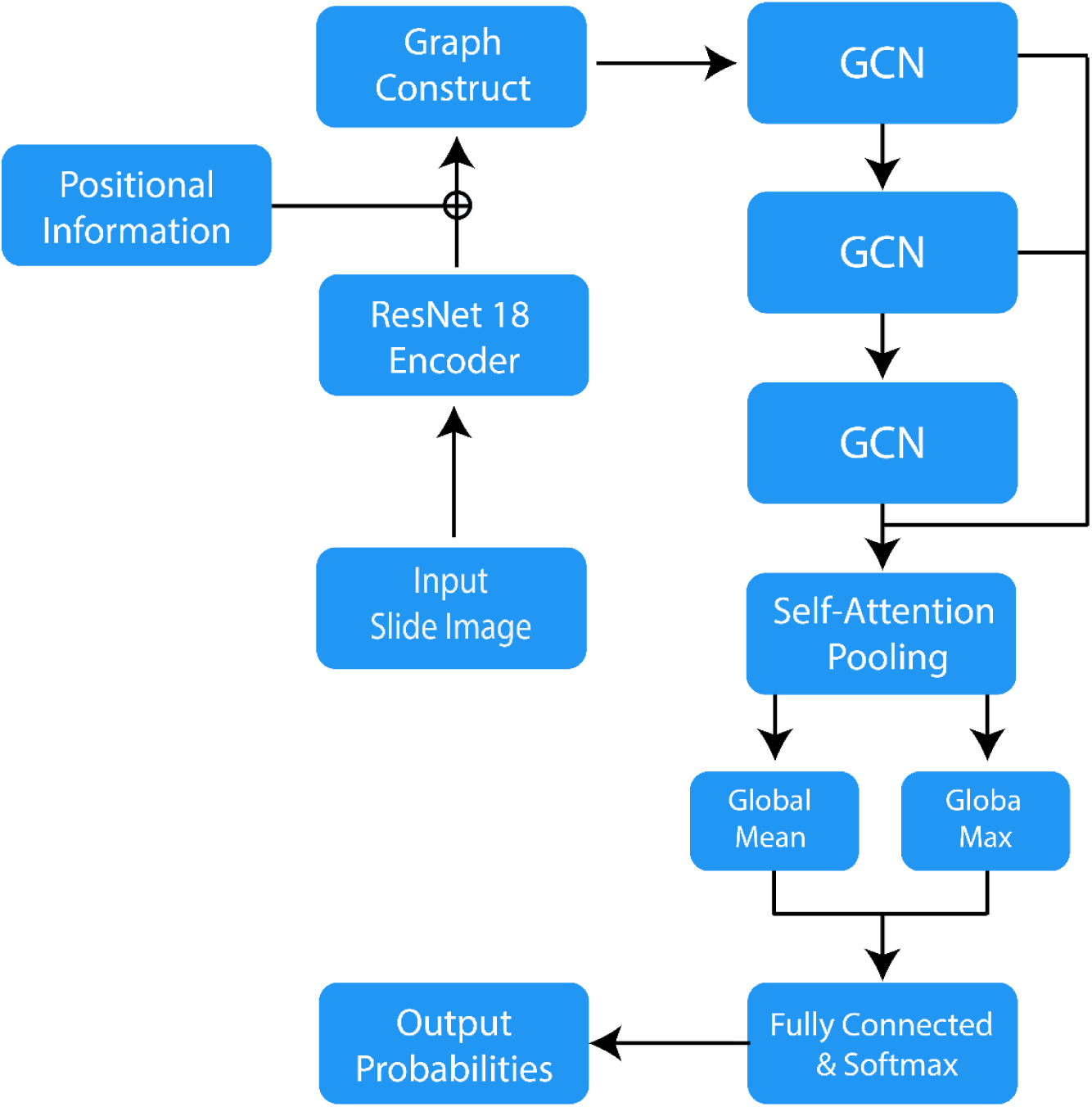
Slide2Graph Architecture

## RESULTS

We evaluated our model’s performance on our holdout test set, which was not used during the model training. **Table 3** summarizes the precision, recall, F1 score, and AUC metrics for each class and overall. In addition, we calculated the 95% confidence intervals for all the metrics using a bootstrapping method with 10,000 iterations. For error analysis, the confusion matrix of our model is shown in **Figure 3**.

**Table 3.** Slide2Graph’s performance metrics and 95% confidence interval on the test set.

|  | Precision | Recall | F1 score | AUC |
| --- | --- | --- | --- | --- |
| Negative | 0.90 (0.67, 1.00) | 0.56 (0.31, 0.80) | 0.69 (0.44, 0.87) | 0.80 (0.66, 0.93) |
| Neoplastic | 0.73 (0.55, 1.00) | 1.00 (1.00, 1.00) | 0.85 (0.71, 1.00) | 0.94 (0.86, 1.00) |
| Positive | 0.79 (0.50, 0.93) | 0.92 (0.73, 1.00) | 0.85 (0.62, 0.95) | 0.90 (0.73, 0.98) |
| Average | 0.81 (0.57, 0.98) | 0.83 (0.68, 0.93) | 0.80 (0.59, 0.94) | 0.88 (0.75, 0.97) |

Abbreviation: AUC, area under the relative operating characteristic curve

**Table 4.** Comparison between Slide2Graph and other baseline models based on bootstrapping and F1 scores and their corresponding p-values.

|  | Slide2Graph | DeepSlide | Decision Tree | Random Forest | Adaboost |
| --- | --- | --- | --- | --- | --- |
| F1 score | 0.79 | 0.70 | 0.64 | 0.70 | 0.72 |
| p-value | - | 7.71e-12 | 4.35e-19 | 4.45e-10 | 3.95e-06 |

**Figure 3.**
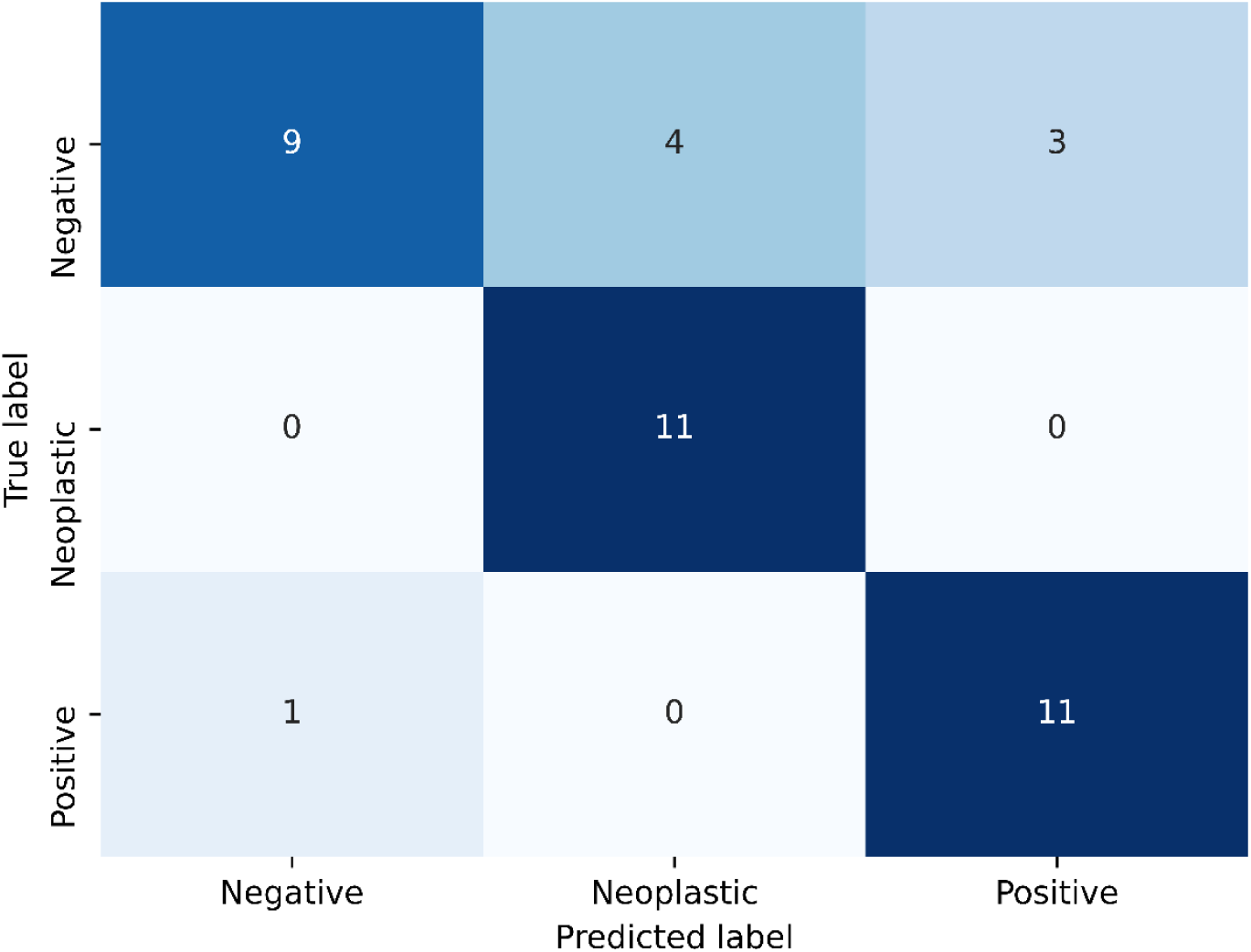
Slide2Graph’s confusion matrix on the test set

### Comparison with other Whole-Slide Inference Methods

We also implemented other models, including DeepSlide, a decision tree, a random forest, and Adaboost ^18,34–36^, to aggregate patch information for comparison and showed the efficacy of Slide2Graph. In DeepSlide ^18,34–36^, we ran systematic grid searches to find the best thresholds for patch-level confidence score and the required percentages of predicted patches in one slide for developing whole-slide inferencing rules using our training and validation sets. In the DeepSlide approach, only patches with a confidence score greater than 0.75 were considered for whole-slide inference. In DeepSlide’s whole-slide inferencing rule, if the percentage of neoplastic or positive patches in a slide exceeded 20% of the entire patches extracted from the whole slide, the slide was classified as neoplastic or positive, respectively. If both the percentages of neoplastic and positive patches exceeded 20%, then the class with the larger percentage was considered as the class for the slide. Otherwise, this slide was deemed to be negative. In addition, we used the percentages of neoplastic and positive patches extracted from a slide as the independent variables to predict the whole-slide label in the other machine learning models, i.e., decision tree, random forest, and Adaboost. Six-fold cross-validation and grid search were used to find the best hyperparameters for the random forest and the Adaboost models. As shown in **Figure 4**, our proposed Slide2Graph graph model performs the best among all five different whole-slide inference models.

**Figure 4.**
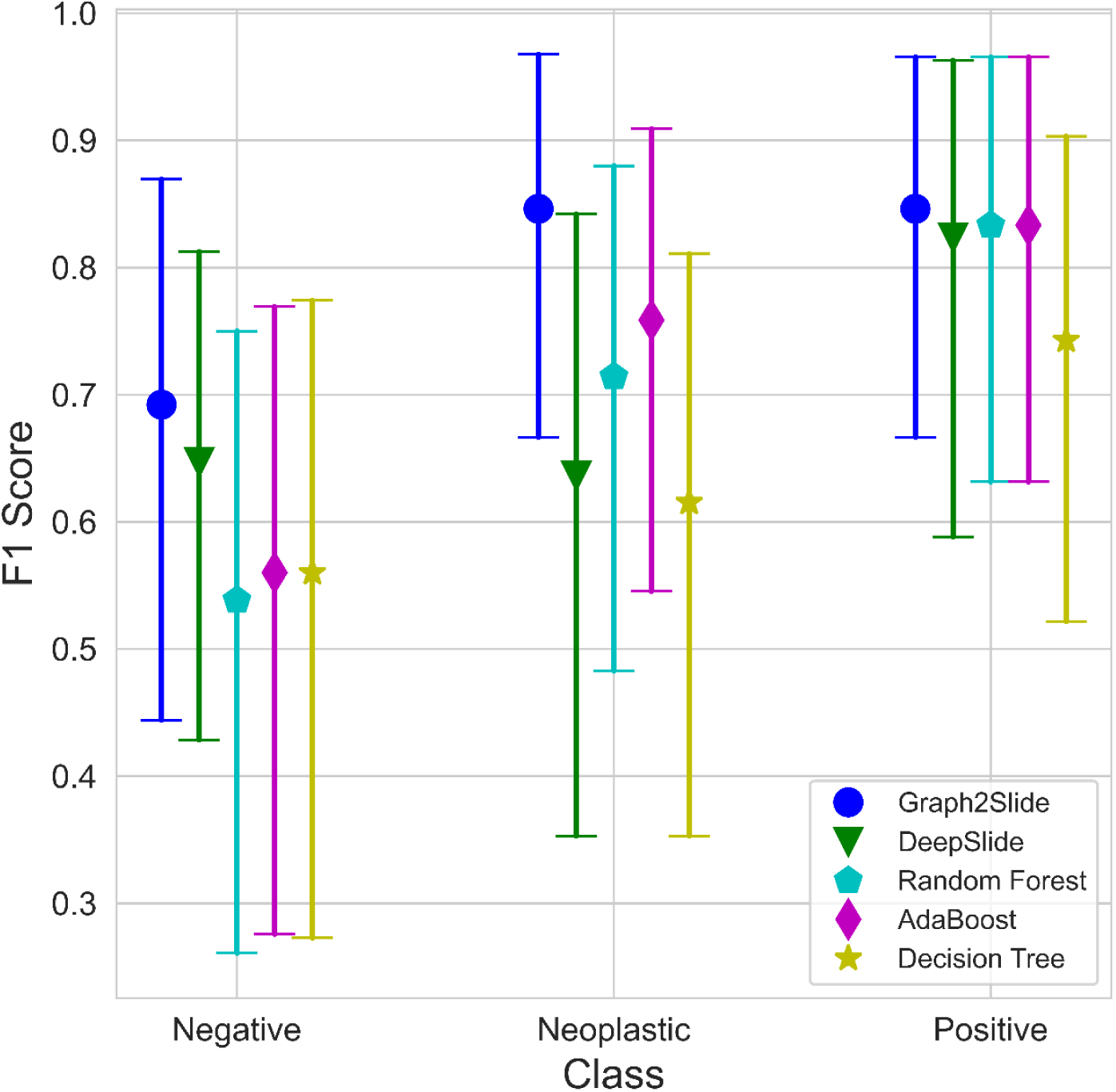
The F1 scores and 95% confidence intervals of different models on the test set stratified by class.

Of note, due to the small size of the testing set, there are overlaps between all models’ F1 score confidence intervals. Therefore, to investigate the statistical significance of the performance difference between different models, we used a bootstrapping approach to randomly sample 50 subsets from our test set. Then, we used the two-tailed student t-test to examine the statistical significance of difference among F1 scores from various methods. As the table below shows, the F1 score of our Slide2Graph model outperformed the other models with a statistical significance level of p-value < 0.0001, while Adaboost was the strongest competitor among the baselines.

## DISCUSSION

The approach proposed in this study can automatically and accurately detect pancreas tumor patterns on the whole-slide images. Our proposed approach achieved the best performance on our test set compared to other baseline methods. Unlike other studies in this domain, which do not consider the benign class in whole-slide inference or rely on detailed benign tissue annotations, we considered tissues outside the region of interest annotations as negative or benign. We used these regions for the training of our patch-level classifier. Therefore, these negative regions could contain noise and findings that may be visually similar to those seen in positive or neoplastic cases. For example, **Figure 5** is an image from a case diagnosed with benign pancreatitis. Albeit the slide is correctly labeled as negative or benign, this region does appear atypical because it contains inflammation and fibrosis surrounding residual atrophic, mild atypical ductular structures (i.e., reactive atypia). This region bears a striking resemblance to a well-differentiated adenocarcinoma with infiltrating glands ^37^. Although the cells in this region are still benign, the tissue overall is architecturally more similar to a well-differentiated adenocarcinoma than to normal pancreatic acini/parenchyma. Likewise, **Figure 6** depicts several situations in which benign pancreas might mimic a neuroendocrine tumor; contrariwise, a neuroendocrine tumor may mimic lymphocytes (if discohesive) or carcinoma (if glandular or organoid)^38^. This phenomenon of mimicry is likely one of the reasons why our patch-level classifier does not perform perfectly, as tissue findings of some reactive and malignant processes are known to demonstrate considerable morphologic overlap ^37,38^. Because we do not explicitly annotate benign (including reactive atypia) or normal regions in our dataset, the developed algorithm is subject to these ambiguities.

**Figure 5.**
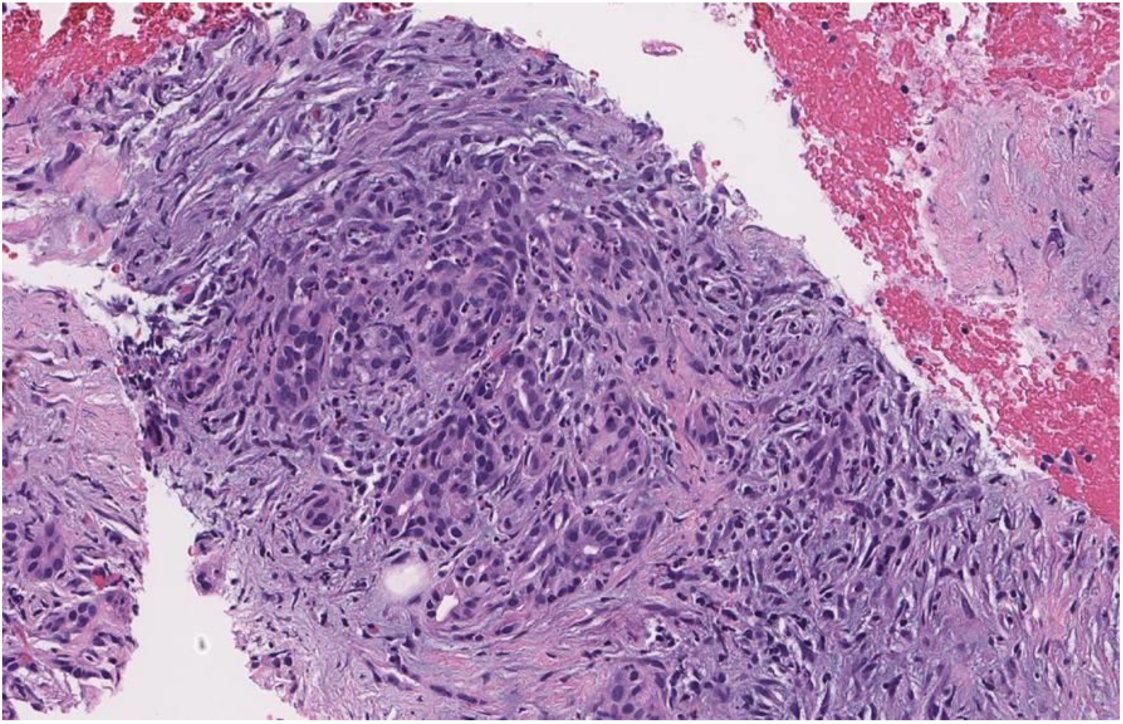
Differential survival of benign pancreatic ducts in chronic pancreatitis can create the illusion of a neoplasm.

**Figure 6.**
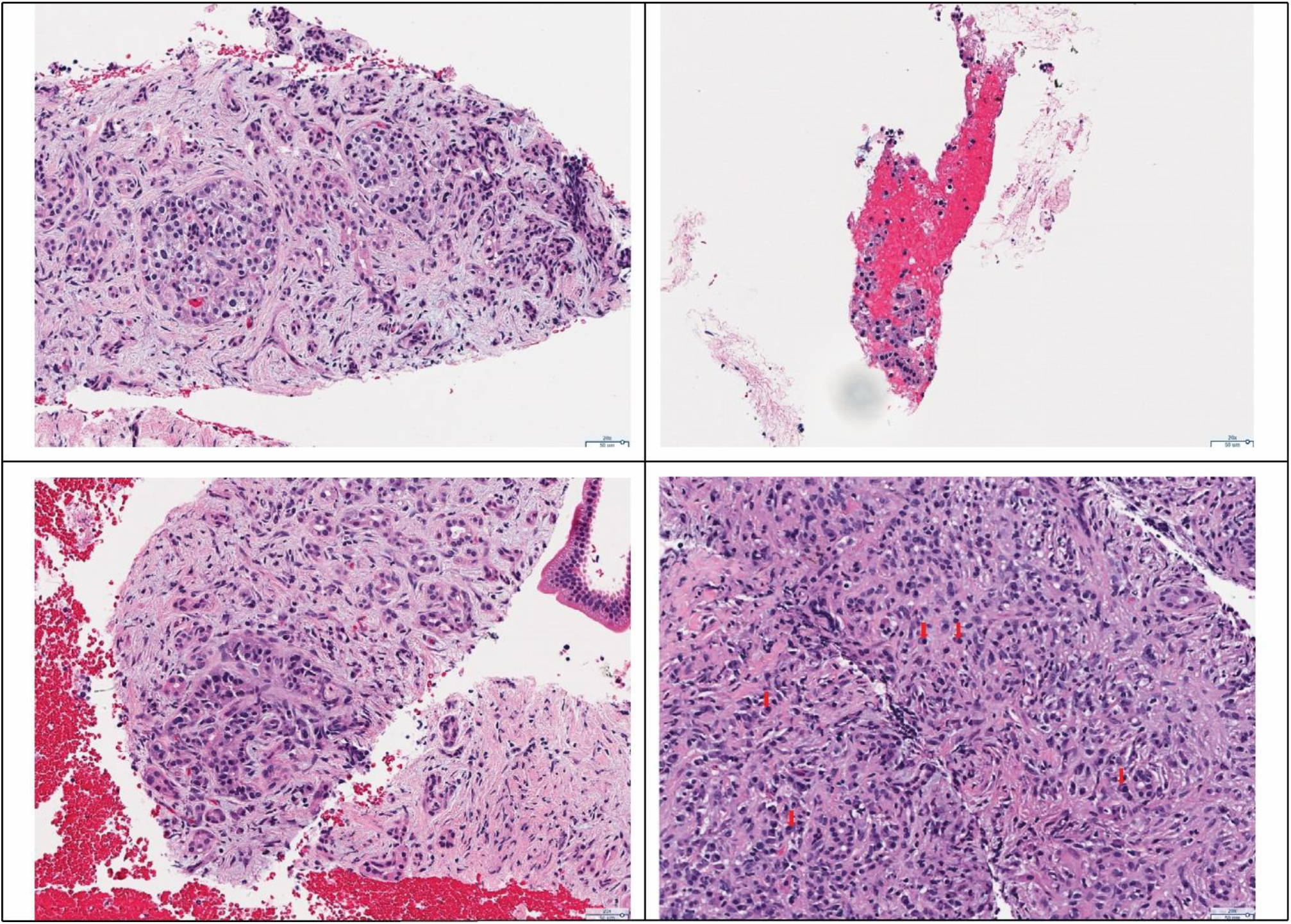
**Top left**: Prominent islet cell aggregates in chronic pancreatitis mimicking neoplastic cells. **Top right**: Detached and degenerated acinar cells may mimic the detached cells of a neuroendocrine neoplasm. **Bottom left:** Surviving ducts in this case of chronic pancreatitis demonstrate an “organoid” pattern, an architecture frequently associated with neuroendocrine tumors. **Bottom right:** Plasma cells may resemble single neoplastic neuroendocrine cells, as seen here in IgG4 related autoimmune pancreatitis.

Notably, although our patch classifier does not achieve the perfect performance at the patch level, our whole-slide inference model still showed a high performance in detecting neoplastic and positive patterns. Reviewing the model’s mistakes by expert pathologists shows that our algorithm’s major error type is “over-calling” of negative cases – i.e., labeling them as either neoplastic or positive. Although this error is diagnostically irritating and would be cumbersome in the full-scale clinical use of the algorithm, it does not create the same concern for patient safety as frequent misclassifications of positive or neoplastic cases as negative.

### Why the Graph Model Performs Better

It is challenging to classify pancreatic tissue in small patches due to significant histologic overlap between low-grade malignancies, reactive atypia, pancreatitis, etc. For example, although **Figure 5** shown above appears benign to trained eyes, it is impossible to exclude the possibility that it might have come from a tissue specimen that contains a tumor; the inflammation and fibrosis seen could easily be found in a case of pancreatitis or at the edge of a malignancy. The ductules seen are minimally atypical and unlikely to be malignant. However, if more atypical epithelium were present elsewhere in the specimen, that evaluation must be reconsidered. Neuroendocrine tumors also often exhibit morphologic overlap with other conditions; **Figure 6** depicts several situations where this may occur. Of note, at our institution, in recent memory, we have received at least one consultation in which prominent islet cell aggregates deceived the initial reviewer into diagnosing a neuroendocrine tumor. However, the evaluation of the subsequent surgical specimen revealed benign chronic pancreatitis as the source of the neoplastic-appearing cells.

Therefore, the patch-level classifier essentially performs as a “weak” classifier due to these overlaps. However, our proposed graph model can consider those patch-level predictions and additional positional data to come to a more reliable prediction for classifying a whole slide, particularly for the neoplastic and positive classes.

Several improvements in our approach facilitate this performance. Unlike the models previously used for whole-slide level inferencing which treat every patch equally and use the percentages of predicted patches for each class to make the whole-slide predictions, our graph-based model takes the patches’ positions and the global structure of the whole slide into consideration. As we applied the self-attention pooling layer to aggregate the node features, we can obtain the associated attention map for a whole slide. This attention map can provide insights into the results of our Slide2Graph model by highlighting the highly weighted regions which significantly influence the whole-slide inference. **Figure 7** shows the highly weighted regions by Slide2Graph in green, which contributed the most to the whole-slide level classification compared to regions of interest annotated by an expert pathologist.

**Figure 7.**
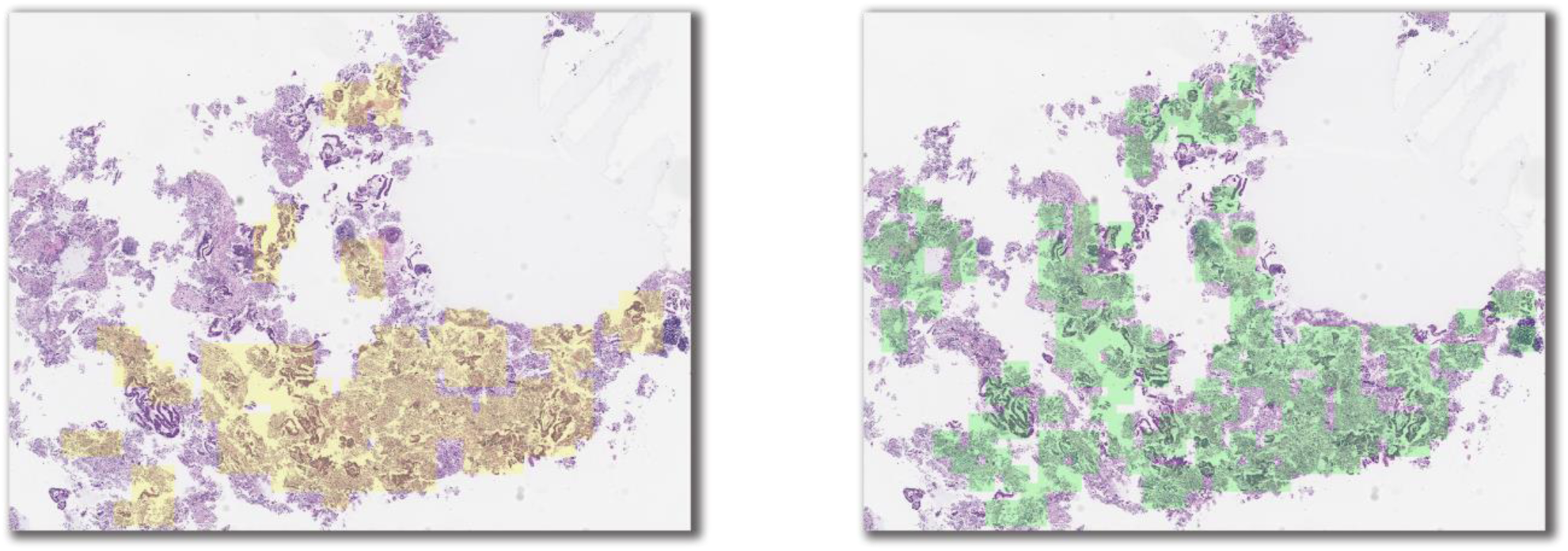
**Left)** Regions of interest annotated by pathologists; **Right)** Highly weighted regions by Slide2Graph for whole-slide inferencing.

### Limitations and Future Directions

The proposed method in this paper has some limitations. First, the size of our dataset in this study is small. Even though we utilize a partially annotated dataset, our sample size, especially in the test and validation sets, is still slim, which resulted in wide 95% confidence intervals in model evaluation.

Moreover, when annotating the whole-slide images, instead of annotating specific negative regions, we only annotated positive and neoplastic regions. As a result, the negative regions used to train our model contain many different types of cells other than positive and neoplastic cells, such as benign epithelium, acinar tissue, normal cells, and inflammation. The variance and diversity of cells in negative regions introduce noise in our negative class and some morphologic overlap with the positive and neoplastic classes. The inclusion of cell blocks from cases overall labeled “atypical” in the negative class may have introduced a low level of additional noise, as well. Although our reviewers considered atypia in the cell blocks not significant, there is some interobserver variability in the threshold at which specimens are called “atypical.” The broad scope of the negative class, while important from the perspective of clinical utility (since it reflects the diversity of negative clinical findings), makes the model harder to train. That is likely why our patch-level classifier did not perform ideally in terms of precision. In future work, we plan to use a larger dataset with a more specific breakdown of annotated negative findings to train our model. In addition, our model is composed of two parts, a patch-level feature extractor, and a whole-slide inference model. These parts are trained separately. Our future work will use small fixed-size patches as nodes directly to construct computational graphs for whole-slide images, so the model can be trained end-to-end. Also, our approach has the potential to perform multi-task learning. We will explore different ways of aggregating the loss function from the patch-level and whole-slide level classifiers to decide whether the information from these two classifiers can benefit each other. Through this process, our model can output the predictions for both patches and whole-slide images simultaneously. In addition, we plan to explore various graph convolutional and global pooling layers and different approaches to construct graphs for whole-slide images to further improve the graph-based model’s performance. Finally, we plan to validate our approach in prospective clinical trials and deploy the developed system as a clinical decision support system in clinical settings.

## Data Availability

All data produced in the present study are available upon reasonable request to the authors.

## ACKNOWLEDGEMENTS

This research was supported in part by grants from the US National Library of Medicine (R01LM012837) and the US National Cancer Institute (R01CA249758).

